# Dorsolateral Prefrontal Cortex Metabolic Profiles in Autism Spectrum Disorder Correlate Atypically with Nonverbal IQ and Typically with Attention Switching

**DOI:** 10.1101/2022.09.17.22280045

**Authors:** Akila Weerasekera, Adrian Ion-M□rgineanu, Nicole M. McGuiggan, Nandita Shetty, Robert M. Joseph, Shantanu Ghosh, Mohamad Alshikho, Martha R. Herbert, Tal Kenet, Eva-Maria Ratai

**Author notes:** **Corresponding Author**: Eva-Maria Ratai, Athinoula A. Martinos Center for Biomedical Imaging, Massachusetts General Hospital, Department of Radiology, Neuroradiology Division, 149 13th Street, Suite 2318, Charlestown, MA 02129, USA. Share senior authorship (Tal Kenet, Eva-Maria Ratai).

## Abstract

**Background:** The neurometabolic profile of associated with autism spectrum disorder (ASD) has been reported to be abnormal by some studies showing region specific metabolite levels in ASD, while others report no group differences. The neurometabolic profile of the left dorsolateral prefrontal cortex (DLPFC) is of particular interest due to the DLPFC relevance to cognitive and executive function, and to ASD.

**Method:** We used ^1^H-MRS to investigate neurometabolic profiles in the DLPFC of ASD and sex/IQ-matched typically developing (TD) children (ages 9-13). We focused on levels of Glutamate and Glutamine (Glx) due to many reported Glx abnormalities ASD, and of Choline (Cho) because of its relationship to intelligence quotient (IQ) and to attentional re-orienting difficulties.

**Results:** While no significant group differences were observed in absolute concentrations, metabolite levels were correlated with the behavioral phenotype of ASD children. In the ASD group but not the TD group, nonverbal IQ (NVIQ) was negatively associated with Cho (r=-0.59, p=0.026) and positively associated with Glx/Cr (r=0.66, p=0.011). Furthermore, attentional-switching scores in the ASD group correlated negatively with Cho (r=-0.69, p=0.009), and positively with Glx/Cr for both the ASD (r=0.73, p=0.004) and TD (r=0.54, p=0.040) groups.

**Conclusions:** Cho and Glx/Cr have different neurometabolic roles in modulating NVIQ in ASD compared to TD children, while their role in attentional switching seems preserved in ASD. Elucidating the apparently divergent role of neurometabolites in ASD in the absence of significant group differences in absolute levels is an important step towards understanding and mapping the neural correlates of ASD. These results are also relevant in the context of the significant cognitive function heterogeneity associated with the ASD phenotype, as they suggest possible underlying neural mechanisms that do not overlap which those expected from typical development.

## 1. INTRODUCTION

Autism spectrum disorder (ASD) is an early onset neurodevelopmental disorder characterized by deficits in social interaction and communication and limited and repetitive behaviors and interests (Margari et al., 2018; Hiremath et al., 2021). Studies of ASD have shown a wide range of abnormalities in brain function and structure (Müller and Fishman, 2018; Forde et al., 2020), as well as abnormalities in the levels of brain metabolites. However, many questions about the underlying neuropathology of ASD remain unanswered (Blatt, 2012; Howlin, 2021).

In this study, we sought to gain a better understanding of how metabolite imbalances in the frontal cortex may contribute to ASD symptomatology. The association of the frontal cortex in the neurobiology of ASD has long been recognized (Courchesne et al., 2011; Stockman, 2013; Stoner et al., 2014). The frontal regions of the brain are known to play a key role in executive and socioemotional function, two of the cognitive processes known to be impaired in ASD (Kim et al., 2015). In particular, structural and metabolic irregularities have been reported in the dorsolateral prefrontal cortex (DLPFC) in ASD, and the DLPFC has long been associated with deficits in executive function and social cognition (Alexander et al., 1986; Haznedar, 2006; Schmitz et al., 2007; Kalbe et al., 2010).

Postmortem studies suggests that children with ASD have an abnormally larger number of neurons in the DLPFC than typical developing children, and that the DLPFC contain areas of immature cells that do not exhibit the regular layered organization of the cerebral cortex (Courchesne et al., 2011; Lainhart and Lange, 2011). Complementing these findings, neuroimaging studies of ASD individuals have shown atypical structural and functional connections between the PFC and other brain areas (García Domínguez et al., 2013; Supekar et al., 2013).

Metabolic abnormalities in ASD have been mapped primarily using postmortem histological methods, as well as non-invasively using ^1^H-magnetic resonance spectroscopy (^1^H-MRS, henceforth MRS), a technique that allows the detection and quantification of absolute and relative concentration of neurometabolites. Postmortem studies of frontal cortex abnormalities in ASD have found GABAergic, glutamatergic, mitochondrial, and microglial dysfunction (Blatt, 2012; Wei et al., 2014; Varghese et al., 2017; Fetit et al., 2021). In parallel, MRS based studies of ASD have shown lower GABA levels and higher glutamate levels in ASD children, in line with postmortem studies. Interestingly, alterations in glutamate or combined glutamate and glutamine (Glx) levels in ASD appear to vary between children and adults, with fewer abnormalities, particularly in GABA levels, detected in adults, suggesting age or disorder associated changes over the lifespan (Naaijen et al., 2015; Ajram et al., 2019).

Many of the metabolic abnormalities documented in the DLPFC in ASD or other related disorders also found associations with behavioral phenotypes. For instance, one study reported a negative association between left DLPFC concentrations of glutamate with perspective taking scores in ASD adults (Montag et al., 2008). In parallel, in electrophysiological studies, cholinergic pathways have been associated with atypical social interaction and behaviors, as well as with ASD symptom severity, orientation of attention and sensory integration (Lam et al., 2006; Orekhova and Stroganova, 2014). Furthermore, a recent ^1^H-MRS study conducted in individuals with generalized anxiety disorder (GAD), a common comorbid diagnosis in ASD, reported left DLPFC Cho/Cr levels inversely predicted IQ and predicted the severity of anxiety in individuals diagnosed with generalized anxiety, but this was not the case in the neurotypical control group, further highlighting the relationship between left DLPFC and cognitive functioning, and underscoring the need for further research into behavioral correlates of choline containing metabolites (Coplan et al., 2018). In combination, these studies points to the importance of additional mapping of the metabolic profile of the DLPFC in determining ASD related cognitive deficits.

Furthermore, categorizing the clinical heterogeneity in children with ASD is essential to the formulation of a framework that could lead to development of new research studies and measurements to capture subgroup differences within the disorder. Deficits in cognitive, emotional, and social behavior often manifest differently across ASD subgroups (ie, verbal/nonverbal, high/low IQ). Much of the research so far have focused on the autism phenotype with less intervention needs, often described as “high functioning”. Less than about 15% of studies have targeted the most comprehensively impacted ASD individuals who are nonverbal or minimally verbal (Jack and A. Pelphrey, 2017). Therefore, better understanding of the neurobiological/chemical underpinnings of how heterogeneity in cognitive and executive function may correlated with biological underpinnings is critical for advancing our understanding of the neural bases that underlie heterogeneity in ASD.

Here, we used ^1^H-MRS to investigate the neurometabolic profiles in the left DLPFC of 14 children with ASD ages 9-13, and 16 age-, sex-, and IQ-matched typically developing children. We focused on two metabolites: Glx due to widely documented abnormalities in Glx in ASD (Cochran et al., 2015; Ajram et al., 2017, 2019), and Cho, because of its previously mapped relationship to IQ in the DLPFC (Barbey et al., 2013; Coplan et al., 2018), and to attention re-orienting difficulties due to deficits in cholinergic arousal system (Deutsch et al., 2010; Anand et al., 2011; Orekhova and Stroganova, 2014). We were interested both in possible differences in absolute levels of these metabolites in ASD, and in any association these metabolites may have with assessments of intellectual and executive function (specifically attention), as well as severity of ASD. More specifically, since IQ and attentional switching have been shown to correlate with metabolites in the DLPFC (Lauber et al., 1996; Smith et al., 2004; Barbey et al., 2013), we hypothesized that DLPFC Cho and Glx levels would show significant associations with these neuropsychological scores and may show distinct association patterns in ASD and TD children.

## 2. METHODS & MATERIALS

### 2.1 Study participants and behavioral assessments

We recruited 17 children with ASD and 17 TD children matched on IQ, age, sex, and handedness, for this study. Of these, the MRS data of 3 of 17 ASD children and 1 of 17 TD children did not pass quality control, as detailed below. The remaining sample therefore consisted of 14 children with ASD and 16 TD children. Parents of the participants provided informed consent according to protocols approved by the MGH Institutional Review Board (IRB). Participant assent was also provided in addition to parent consent for participants aged 14-17. Phenotypic data collected from all participants who passed MRS data quality control are summarized in **Table 1**. The age range was 6–17 and 7–17 years in the TD and ASD groups, respectively, with a mean age of 13.3 and median age of 13. All participants were right-handed, with the exception of two ASD individuals, determined from information collected using the Dean Questionnaire (Piro, 1998). Participants with ASD had a prior clinical diagnosis of ASD and met ASD criteria on the Autism Diagnosis Observation Schedule, Version 2 (ADOS-2) (Lord et al., 2012; Hus et al., 2014) administered by a trained research assistant with inter-rater reliability. The Social Communication Questionnaire - Lifetime Version (SCQ Lifetime) (Rutter Bailey, A., & Lord, C., 2003) was administered to further confirm ASD in ASD participants and rule out ASD in TD participants. ASD participants who did not meet a cutoff of ≥15 on the SCQ or who had a borderline score on the ADOS-2 were further evaluated by expert clinician and co-author Dr. Robert Joseph to confirm the ASD diagnosis. Individuals with autism-related medical conditions (e.g., Fragile-X syndrome, tuberous sclerosis) and other known risk factors (e.g., gestation <36 weeks) were excluded from the study. All TD participants were below the threshold on the SCQ Lifetime questionnaire. Parent-questionnaires were administered to confirm that participants were free of any neurological or psychiatric conditions and substance use in the past 6 months. For ASD, the Social Responsiveness Scale (SRS-2) was used to assess the severity of the ASD symptoms (Bruni, 2014). Verbal IQ (VIQ) and nonverbal IQ (NVIQ) were assessed using the Differential Ability Scales – II (Beran, 2007) for all participants. The TD and ASD groups did not differ on VIQ, and showed a non-statistically significant trend towards a group difference on NVIQ. Lastly, all participants completed the INN (Inhibition-Naming), INI (Inhibition-Inhibition), and INS (Inhibition-Switching) sections of the NEPSY-II neurocognitive evaluation. Derived from these sections, the Inhibition Contrast Scaled Score (ICS-I), which measures the ability to voluntarily inhibit attention, and the Switching Contrast Scaled Score (ICS-S), which measures the ability for switching attention between competing stimuli, were computed for each participant.

**Table 1.**
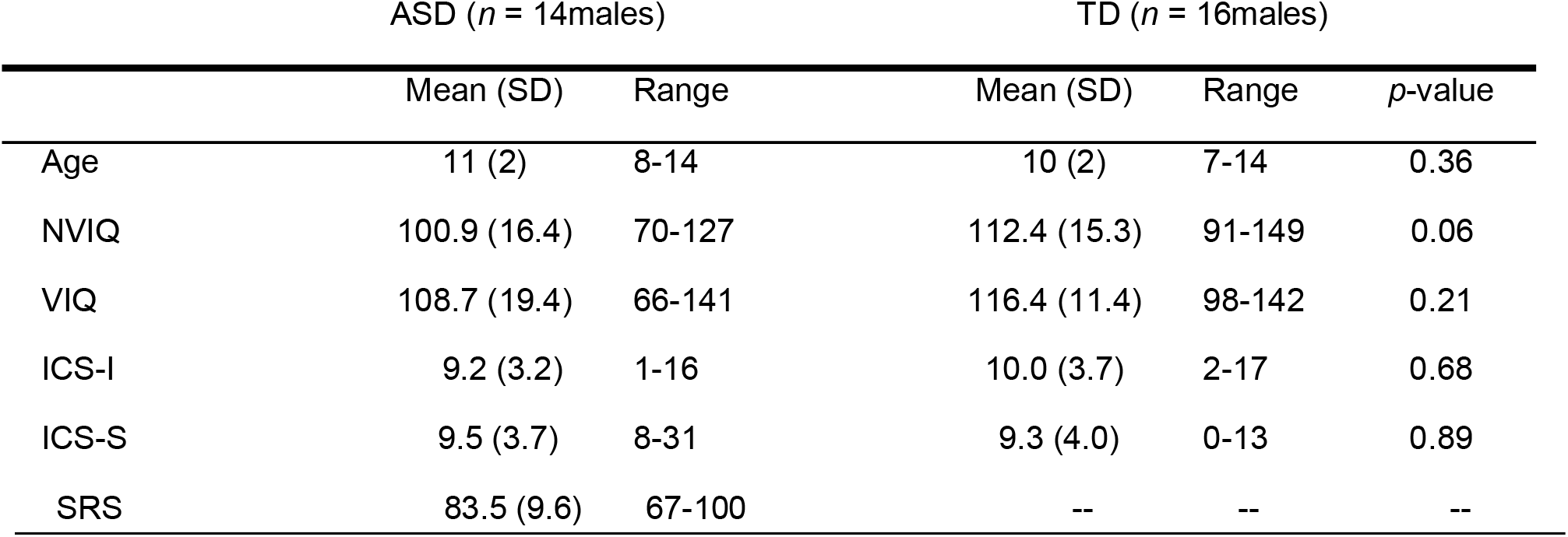
Characterization of the participants.

The *p*-values are from two-sample *t*-tests for the difference in means between the ASD and TD groups. NVIQ: nonverbal IQ; VIQ: verbal IQ; ICS-I: inhibition of attention, as measures on the Nepsy-II; ICS-S: attentional switching as measures on the Nepsy-II; SRS: Social Responsiveness Scale.

### 2.2 Brain imaging data acquisition and processing

Brain imaging was performed using a 3T Siemens Trio MR scanner (Siemens Healthineers, Erlangen Germany) equipped with a 12-channel head coil. In all participants, a high-resolution multi-echo Magnetization Prepared - RApid Gradient Echo MEMPRAGE (T1-weighted structural MRI) volume was also acquired (TR/TE1/TE2/TE3/TE4 = 2530/1.69/3.55/5.41/7.27 ms, flip angle = 7°, voxel size = 1 mm isotropic), for the purpose of anatomical localization, MRS voxel placement and the correction for partial volume effects of cerebrospinal fluid (CSF).

^1^H-MRS protocol: Single voxel MRS was acquired using a conventional Point RESolved Spectroscopy (PRESS) sequence (TE=30ms, TR=2.5s, bandwidth=1.2 kHz, and 96 averages, 1024 sample points). A 20×20×20 mm voxel was placed in the left DLPFC (**Figure 1A**).

**Figure 1:**
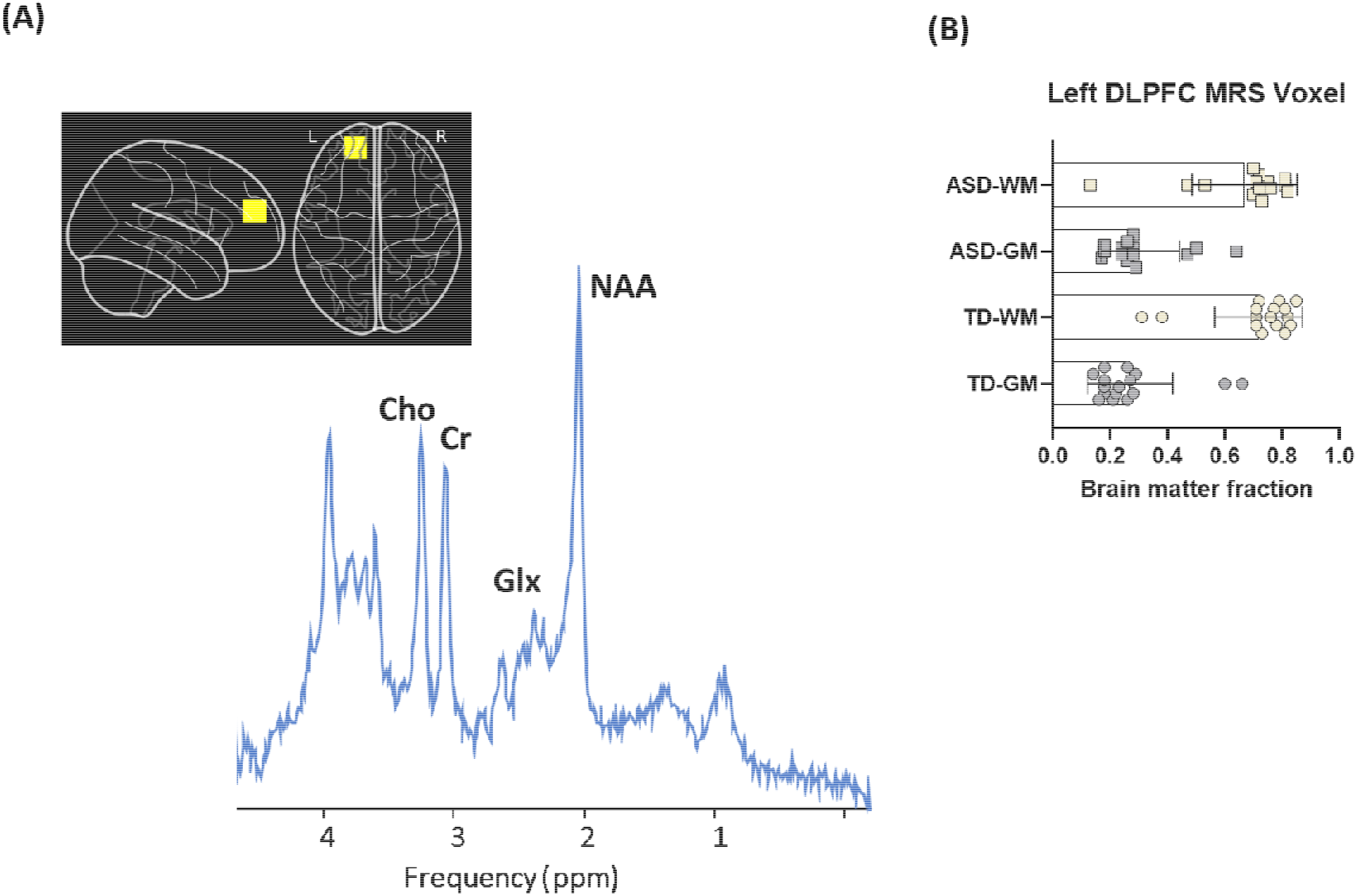
Voxel placement, representative spectrum and brain matter fractions. **(A)** Voxel placement on left DLPFC and representative spectrum. (**B**) Gray and white matter (GM, WM) fractions within the 8 cm^3^ left DLPFC-voxel in ASD and TD subjects.

MR spectra were processed off-line using the LCModel software, version 6.3 (Provencher, 2001) for quantitative assessment of the following neurometabolites: N-acetylaspartate (NAA), Creatine (Cr), choline (Cho), myo-inositol (mI), and glutamate+glutamine (Glu+Gln). LCModel analysis was conducted on spectra within the chemical shift range 0.5–4.1 ppm. Spectra were excluded when the signal to noise ratio (SNR), estimated by LCModel (defined as peak height of NAA divided by the root mean square of the noise of the LCModel fit) was less than 5 and the Cramér-Rao lower bounds (CRLB) higher than 20%.

All metabolite levels were adjusted for gray matter (GM), white matter (WM), and cerebrospinal fluid (CSF) contributions as follows: the MEMPRAGE images and the voxel coordinates from the Siemens RDA files were used in Gannet toolkit version 3.1 (Edden et al., 2014) to generate binary masks of the voxel location. These masks were then used in SPM 12 (Penny et al., 2007) to calculate the partial volumes of GM, WM and CSF percentages within the voxel. The segmented tissue fractions were then used to correct for metabolite concentrations quantified using LC Model for CSF content according to the literature (Gasparovic et al., 2006).

### 2.5. Statistical analyses and multiple comparisons

Statistical analysis was performed using Prism GraphPad v9 (GraphPad, La Jolla, California) with unpaired, two-tailed t-tests on the ASD and TD groups. Pearson correlation coefficients were calculated to assess the relationship between neurometabolite concentrations and each behavioral measure. Additionally, we assessed effects of combined metabolite levels in ASD and TD with a multivariate linear regression model in GraphPad (least squares method: Y_Behavior_= β0 + β1[met1]_group_ + β2[met2]_group_, where dependent variable Y is the behavior measure and continuous predictor variables are metabolite levels).

We tested for group differences in ASD versus TD in mean concentrations of Cho, Glx, Cr, for 3 tests in all; no significant group differences emerged, and so we did not correct the results for multiple comparisons. We also tested the following correlations within each group: Cho correlations with VIQ, NVIQ, SRS (ASD only), ICS-S, ICS-I, and Glx/Cr with the same 5 behavioral metrics, for 10 correlations tests in total in the ASD group, and 8 in the TD group. Lastly, we tested for significant differences between slopes for the ASD vs TD groups only for correlations between Cho and NVIQ, and Glx/Cr and NVIQ.

When considering a correction for multiple comparisons, we first considered potential dependencies across the behavioral measures. As expected, NVIQ and VIQ were significantly correlated (combined groups, r=0.52, p=0.003), and NVIQ and ICS-S were also significantly correlated (combined groups, r=0.53, p=0.004) ICS-I and SRS scores were not correlated with ICS-S, ICS-I, VIQ or NVIQ. Because the behavioral measures are not statistically independent, it is challenging to account for such dependencies when correcting for multiple comparisons.

We therefore chose to report uncorrected p-values, following recent best practices (Lu and Belitskaya-Levy, 2015; Amrhein et al., 2019; Lowe, 2019). We used * to mark p values of 0.05 or below, and ** to mark p-values of 0.005 or below, i.e. p values that would survive a Bonferroni correction for 10 comparisons.

## 3. RESULTS

### 3.1 Tissue composition within the MRS voxel

We began by testing whether the voxels chosen across the two groups had similar compositions. As expected, the groups did not differ significantly in mean grey matter, white matter, or CSF fraction within the voxel (**Figure 1B**) (p > 0.05 for all).

### 3.2 Neurometabolite levels and age effects

Next, we investigated group differences in absolute metabolite levels, for Glx and Cho. No significant differences in absolute levels of these two neurometabolites were observed between groups (**Figures 2A-B**, and **Table 2**). Because maturation trajectories of metabolites in this age range are not well understood (Nelson et al., 2019; Porges et al., 2021), we then tested for age effects of metabolite concentrations within our sample. No significant age-related metabolite changes were observed for either group (p>0.05) for Cho (**Figure 2D**), while a negative trend was observed for Glx in the ASD group (r= -0.53, p=0.052) (**Figure 2E**). Given this trend, we opted to normalize Glx levels with Creatine (Cr) levels, as is common practice (Wilson et al., 2019). The Cr referenced Glx levels still did not differ between groups (**Figure 2C**), and the trend with age in the ASD group was indeed removed by this normalization (**Figure 2F**).

**Figure 2:**
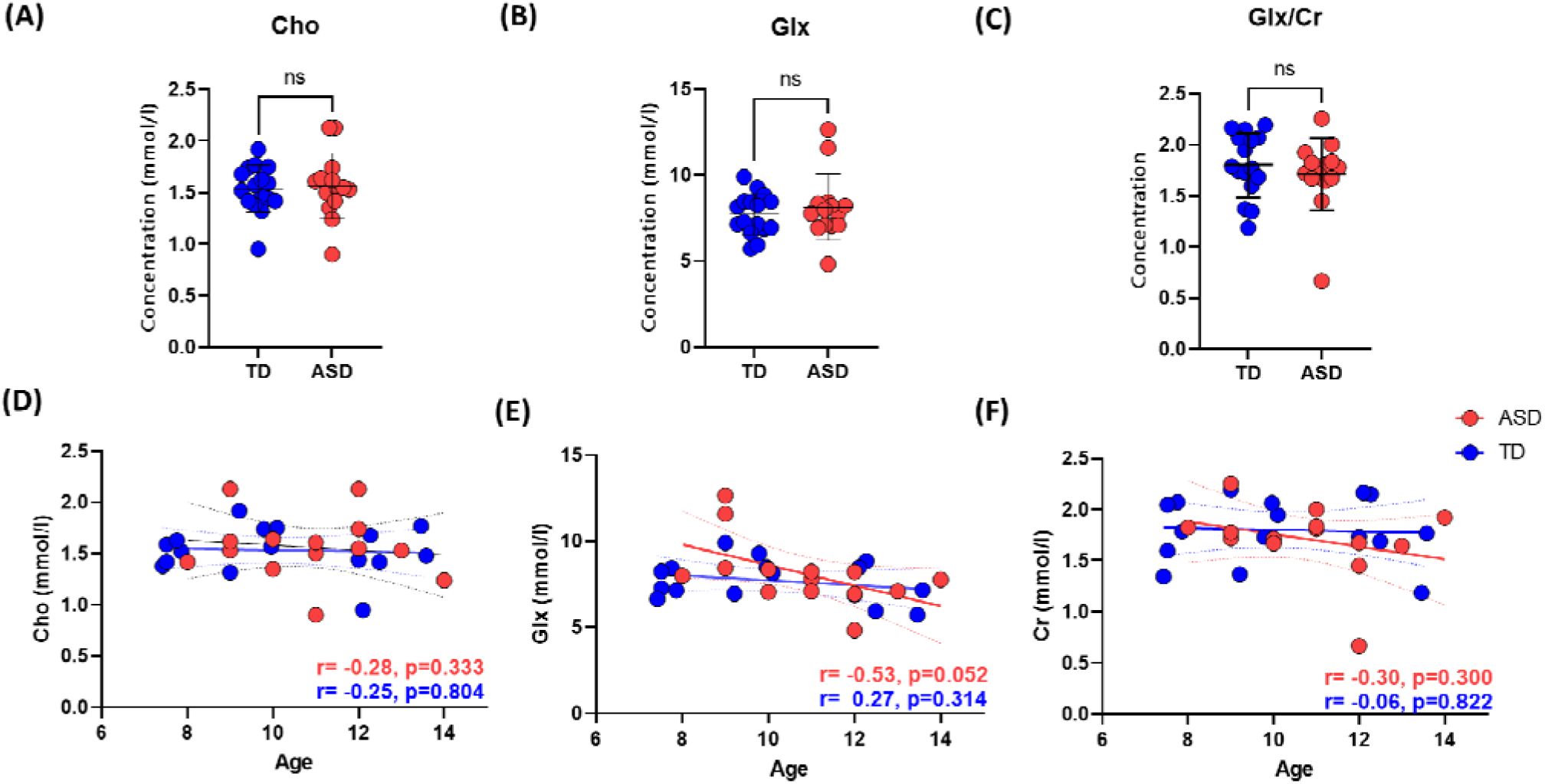
Metabolite concentrations between groups, and correlation with age. Top Row: Group comparisons for **(A)** Cho, **(B)** Glx, **(C)** Glx/Cr. Bottom row: Correlations between age and metabolites levels for **(D)** Cho, **(E)** Glx, **(F)** Glx/Cr.

**Table 2:**
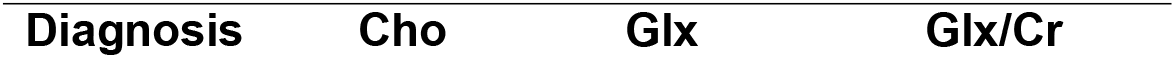
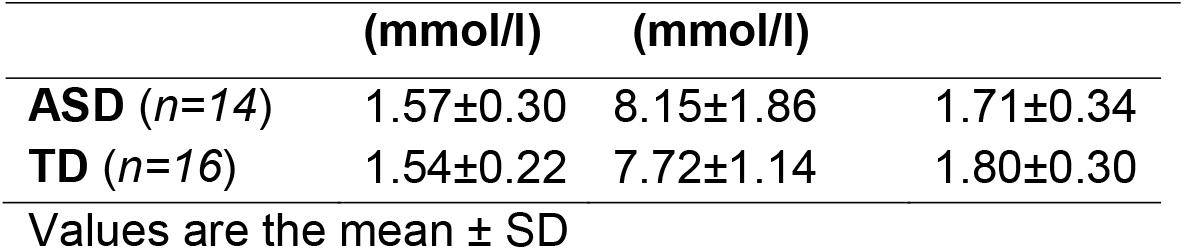
Average metabolite levels

### 3.3 Correlations between Cho and behavioral measures

We then investigated the relationship between Cho levels in the left DLPFC and VIQ/NVIQ, ASD severity, and measures of attention. Between VIQ and NVIQ, significant correlations were found only with NVIQ (**Figure 3A**). In the ASD group, absolute Cho levels showed a negative correlation with NVIQ (r= -0.59, p=0.026). In contrast, in the TD group, Cho levels correlated positively with NVIQ (r= -0.68, p=0.004). There was a significant group difference in the slopes, indicating a significant group difference in the interaction between NVIQ and Cho.

**Figure 3:**
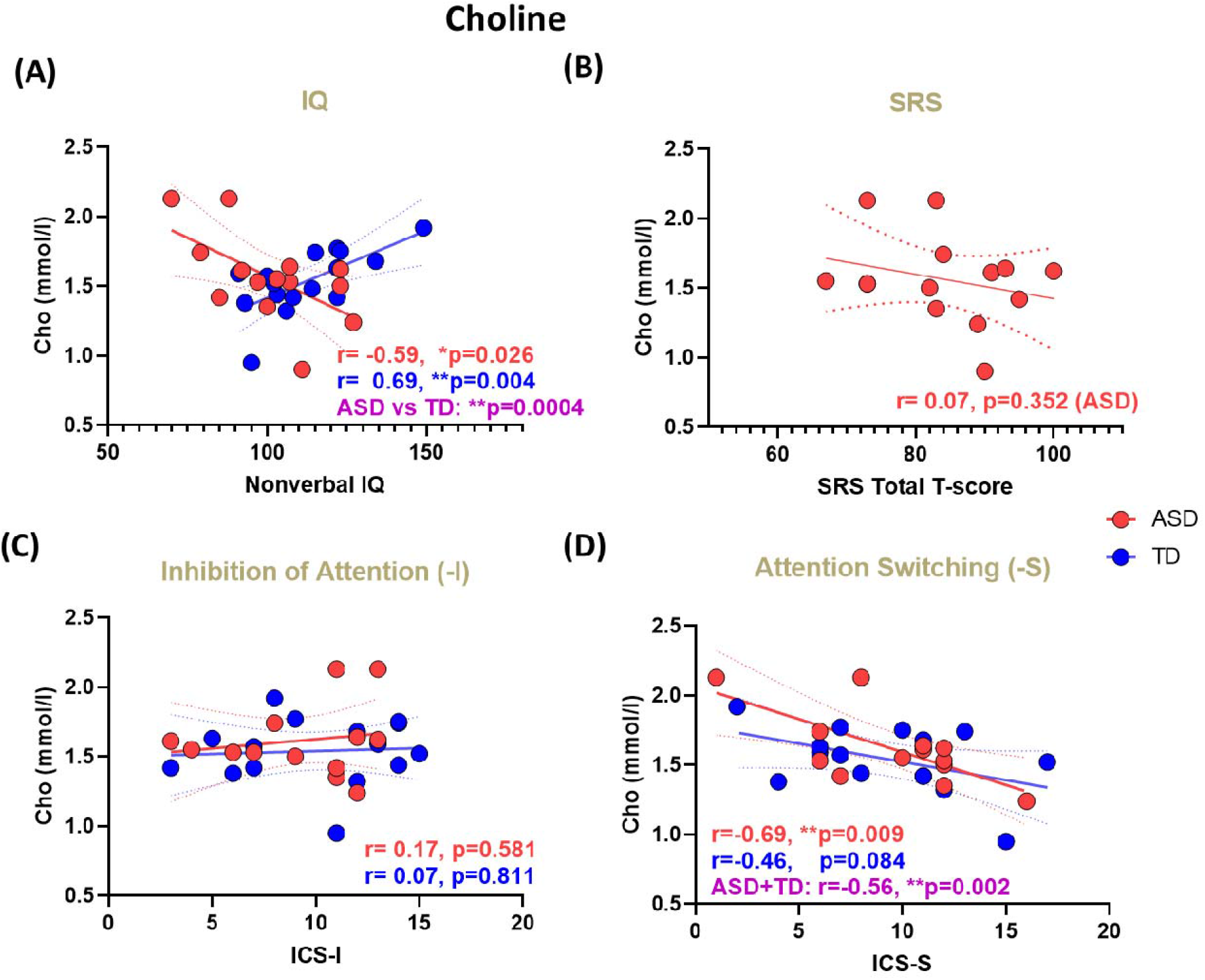
Correlation between behavioral measures, and Cho. **(A)** NVIQ. r and p values are shown for each group separately. A statistical ASD vs TD comparison of difference in slopes shows a highly significant difference between the two slopes (purple) **(B)** SRS. **(C)** inhibition of attention. **(D)** Attentional switching. Since both groups showed a similar trend, we also computed r and p values for both groups combined (purple). **Red:** ASD. **Blue:** TD. Dashed lines indicate the 95% confidence intervals for regression lines. r=correlation coefficient (Pearson’s). p=uncorrected p-values.

No significant correlations were found between Cho and ASD SRS scores, which assess ASD severity (**Figure 3B**).

Lastly, we looked at two behavioral measures of attention: inhibition of attention (ICS-I) and attentional switching (ICS-S). While no significant correlations were found between ICS-I and Cho (**Figure 3C**), ICS-S values in both groups correlated significantly with Cho (**Figure 3D**), and this significant correlation was especially pronounced in the ASD group (r=-0.69, p=0.009).

There was no group difference in the direction of this correlation, and indeed, combining the two groups strengthened the correlation (r=-0.56, p=0.002).

### 3.4 Correlations between Glx/Cr and behavior measures

The same analyses were then repeated with Glx/Cr instead of Cho (Section 3.3). As with Cho, VIQ did not correlate with Glx/Cr in either group. As with Cho, Glx/Cr was significantly correlated with NVIQ in the ASD group, albeit in the opposite direction – positively rather than negatively; in contrast to the finding with Cho, there was no significant correlation between Glx/Cr and NVIQ in the TD group (**Figure 4A**).

**Figure 4:**
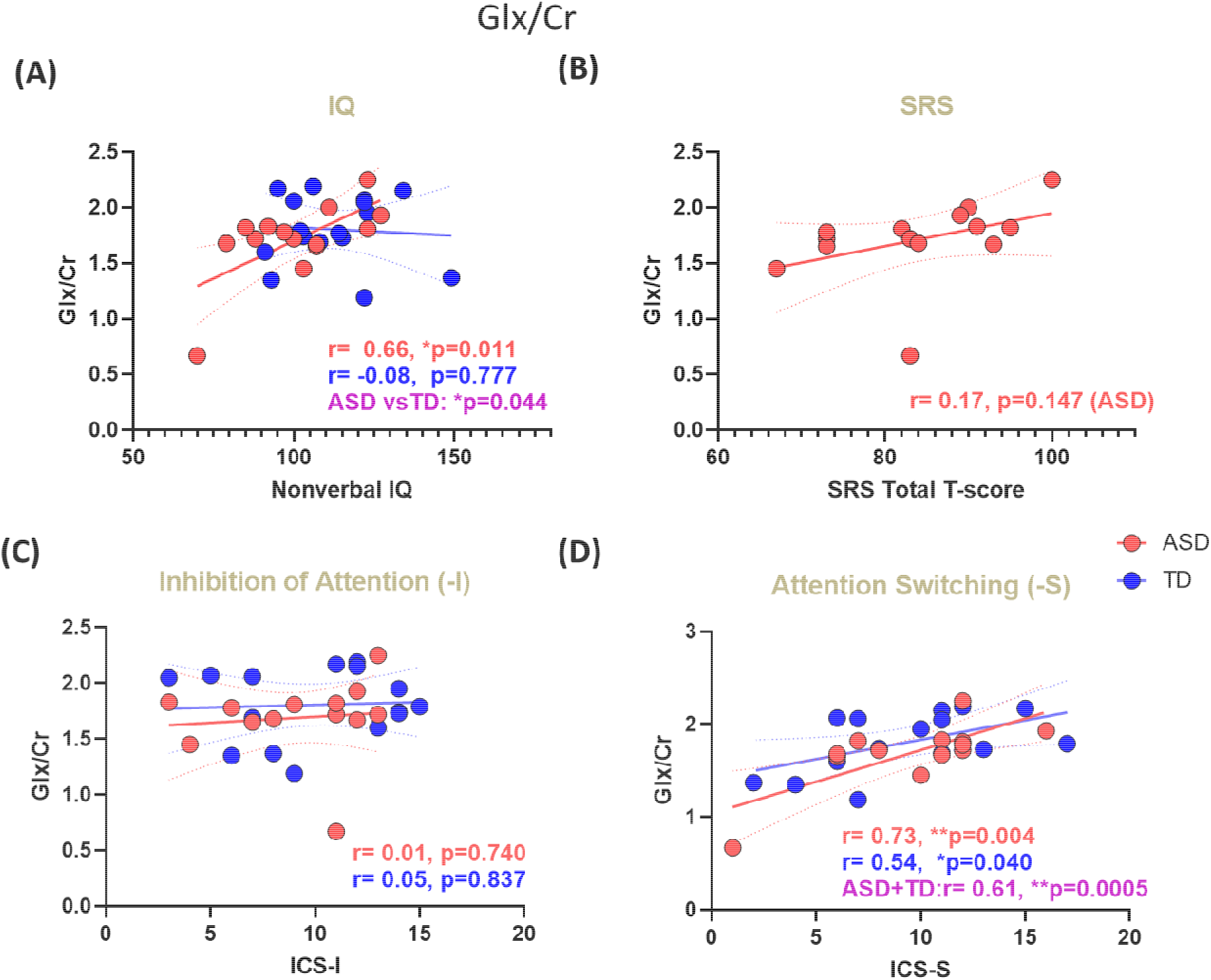
Correlation between behavioral measures, and Glx/Cr. **(A)** NVIQ. r and p values are shown for each group separately. A statistical ASD vs TD comparison of difference in slopes shows a highly significant difference between the two slopes (purple) **(B)** SRS. **(C)** inhibition of attention. **(D)** Attentional switching. Since both groups showed a similar trend, we also computed r and p values for both groups combined (purple). **Red:** ASD. **Blue:** TD. Dashed lines indicate the 95% confidence intervals for regression lines. r=correlation coefficient (Pearson’s). p=uncorrected p-values.

No significant association was found between Glx/Cr levels and the SRS scores (ASD group) (**Figure 4B**). Glx/Cr did not correlate with ICS-I (**Figure 4C**), and was positively correlated with attentional switching (**Figure 4D**) for both ASD (r=0.73, p=0.004) and TD (r=0.54, p=0.040) groups. Indeed, when the ASD and TD groups were combined, we again observed an increased statistical significance for the correlation between ICS-S and Glx/Cr (r=0.61, p=0.0005).

Lastly, a multivariate linear regression model: Y_[Beh]_ ∽ Intercept (β0) + β1[Glx/Cr]_[TD/ASD]_ + β2 [Cho]_[TD/ASD]_, showed that the combined Cho and Glx/Cr predicted both behavioral measures, NVIQ (r=0.70, p=0.023) and ICS-S (r=0.80, p=0.005) among ASD children and NVIQ (r=0.72, p=0.009) among TD children (**Figure 5**).

**Figure 5.**
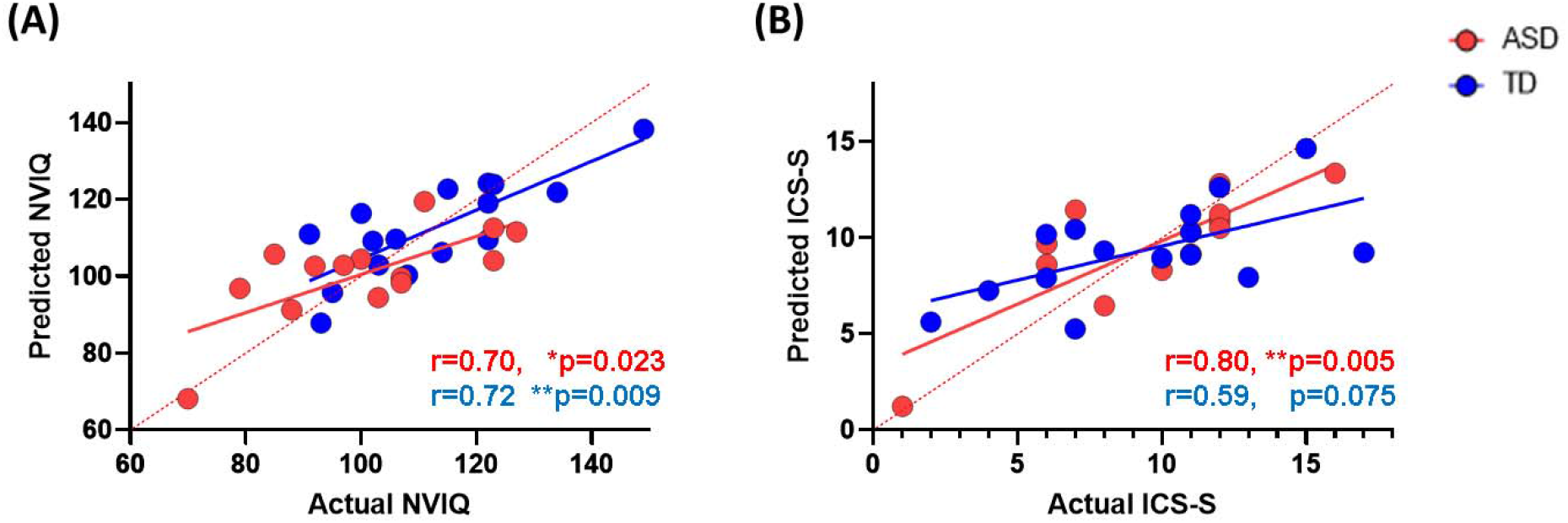
Multivariate linear regression model in ASD and TD. Performance of combined Cho+Glx/Cr predicting **(A)** NVIQ and **(B)** ICS-S

## 4. DISCUSSION

In this study, no significant differences in the left DLPFC metabolite levels, either absolute or creatine referenced, were found between children with ASD and TD children. In spite of no differences in absolute levels of neurometabolites, in agreement with the initial hypotheses, we found significant associations between neuropsychological profiles and metabolites of interest. Specifically, we observed a significant negative correlation between Cho and NVIQ for children with ASD versus a significant positive correlation between these values for the TD group. Choline also showed a significant negative correlation with the ICS-S score (attentional switching) within the ASD group. Even though this association was not significant for the TD group, combining both groups yielded stronger negative statistical significance, meaning both groups followed a similar trend for this association. NVIQ was also positively correlated with the Glx/Cr ratio in the TD group, but not in the ASD group. Additionally, the combination of Cho and Glx/Cr predicted both NVIQ and ICS-S in the ASD group, but only NVIQ in the TD group.

Choline containing compounds are key components of cell membranes, and essential for the synthesis of acetylcholine (ACh), a neurotransmitter linked with learning, memory and attention, that has also been associated with cognitive dysfunction and anxiety severity (Ferguson et al., 2003; Moon and Jeong, 2015). Additionally, Cho levels are also associated with the dopaminergic neuronal transduction and synaptic activity and overall dopaminergic and cholinergic balance, involved in attention, memory, and academic achievements (Wiguna et al., 2012). A previous study of individuals with generalized anxiety disorder reported an inverse association with the left DLPFC Cho/Cr and full scale IQ (FSIQ), while no such relationship was observed for the control group (Coplan et al., 2018). Furthermore, Cho levels have also been associated with white matter density and speculated to reflect excessive neuronal connectivity or abnormal myelination (Laycock et al., 2008). Our finding of a significant association between low Cho levels and higher NVIQ and better performance on attentional switching in ASD is consistent with the classical view. Increased Cho may be a marker of incomplete myelination or neuroinflammation, which is also in part consistent with the prior studies (Jung et al., 1999; Coplan et al., 2018) that found lower Cho levels were associated with increased IQ, and higher Cho levels were associated with increased anxiety Index. This is in contrast to the TD group which is likely not to have any markers of neuroinflammation, and thus in TD, Cho is associated with the synthesis of acetylcholine (ACh), a neurotransmitter associated with memory, learning and attention.

In parallel to the findings of associations with Cho levels, we also found a significant positive correlation between Glx/Cr ratio and NVIQ in the ASD group, but not in the TD group. Glx/Cr ratio also showed a positive association with the ICS-S score for attention switching for both ASD and TD groups, and the combination of the two groups showed a much stronger statistical significance. Glutamate is synthesized from glutamine in neurons and released into the synapse, which is converted back into glutamine in glial cells, by the enzyme glutamine-synthetase. Glutamate to GABA conversion is catalyzed by the neuronal enzyme glutamate decarboxylase. Glutamate and GABA have been found as reliable markers of cortical excitability and inhibition, and thus critical for the mechanisms of neuroplasticity and learning. Moreover, brain excitation and inhibition levels are thought to be critical for triggering the onset of sensitive periods for cognitive skill acquisition by shaping plastic responsiveness of underlying neural systems in response to environmental stimulation (Werker and Hensch, 2015). MRS studies of Glx in neurodevelopmental disorders, including ASD and ADHD, have reported abnormal glutamate or Glx levels in patient populations relative to typically developing subjects (Carrey et al., 2007; Brown et al., 2013; Cochran et al., 2015). Higher levels of glutamate have been hypothesized to indicate hyperexcitability in these disorders that affect neuronal-network dynamics involved in learning, and memory consolidation (Pugh et al., 2014). No group-level differences were found in Glx or Glx/Cr in our study. Yet, we found a significant positive correlation between Glx/Cr and nonverbal intelligence only in the ASD group. This may suggest that DLPFC Glx in ASD may have a different glutamatergic transmission than that of TD children. It also suggests a possible neural mechanism within the DLPFC for the heterogeneity observed in cognitive function in ASD.

There is a multitude of evidence suggesting a neurochemical basis of higher-level cognitive skills, yet the exact mechanisms remain largely unknown. Across developmental disorders such as ASD and attention-related disorders, neurometabolites profiles has been found to vary compared to typically developing age-matched counterparts (Perlov et al., 2009; Baruth et al., 2013). In a ^1^H-MRS study using children with reading disabilities (RD), it was shown that RD children had elevated Cho and glutamate relative to TD children. They further indicated that Cho and glutamate levels inversely correlated with reading and related language measures such that increased concentrations were associated with poorer performance (Pugh et al., 2014). A more recent RD study reported a negative association between Cho and reading ability of children aged between 6 and 8 (Del Tufo et al., 2018). The same study also observed a positive relationship between glutamate concentration and reading performance. Lastly, the fact that in both the ASD and TD groups both Cho and Glx/Cr levels were correlated with attentional switching scores, but not with attentional inhibition, is consistent with the putative role of the prefrontal cortex in attention, since its role for attentional switching, but not inhibition of attention, is well established (Rossi et al., 2009). Attentional switching happens in the twin anticorrelated functional networks involving the DLPFC as a task-positive functional site, demonstrated by Fox and colleagues (Fox et al., 2005). The task-switching aspect and its relationship with neurometabolites in ASD is worth exploring as a biomarker for further investigation. Several limitations of the present study should be noted. Our final analysis included just 14 children with ASD and 16 TD children. Future studies with larger samples are needed to gain greater statistical power and validate our findings. Since we included males between the ages of 7-14 years, our results in this study are representative for only males in this age group. Furthermore, the cross-sectional design of our study limited the power to detect age-driven changes in the neurometabolic profile which could hinder ability to identify disorder specific biomarkers in ASD. Therefore, longitudinal MRS studies are clearly necessary. Despite these limitations it should be noted that it is unlikely that our findings can be fully explained by potential confounds, such as differences in voxel tissue composition, age- or IQ. There were no significant between-group differences in voxel brain matter and CSF, metabolite concentrations were tissue corrected, and the results are consistent with prior metabolites studies of ASD and of other related disorders.

In summary, the main findings revealed no significant group difference in Cho or Glx/Cr metabolite levels between groups, alongside relevant correlations with behavioral measures. Specifically, we documented an inverse relationship between both Cho and Glx/Cr and nonverbal intelligence in ASD children, which was opposite in direction when compared with age-, IQ-matched TD children. This suggests that Cho and Glx/Cr may have different neurometabolic roles in ASD compared to TD children in overall cognitive function. In contrast, for both groups, attention switching scores showed an inverse relationship with Cho and a positive correlation with Glx/Cr, suggesting further that the roles of metabolites for executive function skills, and specifically attentional switching and inhibition of attention, are preserved in ASD. These results are especially interesting in the context of the significant heterogeneity associated with the ASD phenotype (Lenroot and Yeung, 2013), since they suggest possible underlying neural mechanisms for the cognitive heterogeneity in particular, which do not overlap which those expected from typical development. Although the mechanisms connecting these neurometabolites to atypical cognitive functioning alongside typical executive functioning remain to be elucidated, the overall finding that the role of neurometabolites in ASD does not follow a consistent pattern relative to TD children may provide valuable information on potential neurobiological pathways of atypical development, and the heterogeneities associated with cognitive function in ASD in particular.

## Data Availability

All data produced in the present study are available upon reasonable request to the authors

## ACKNOWLEDGMENT

We like to thank our participants and their families. This study was supported by the following: The National Institutes of Health (R01NS048455, MH); the Nancy Lurie Marks Foundation (MH); MIT-MGH Strategic Partnership grant from the Executive Committee on Research (ECOR) at Massachusetts General Hospital (EMR); the National Institute of Child Health and Development (R01HD073254, TK); the National Institute of Mental Health (R01MH117998, TK)

## Author contributions

**EMR, TK, MRH, AW:** Conceptualization; **EMR, MRH, NMM, TK**: Data curation; **AW, AIM**: Data analysis; **EMR, TK, MRH**: Methodology; Project administration; Supervision; **EMR, TK, AW**: Validation; Visualization; **Writing:** AK, EMR and TK wrote the manuscript, all other authors reviewed and or edited the manuscript.

